# Can antiepileptic drug efficacy be studied from electronic health records? A review of current approaches

**DOI:** 10.1101/2020.07.06.20147397

**Authors:** Barbara M Decker, Chloé E Hill, Steven N Baldassano, Pouya Khankhanian

## Abstract

As automated data extraction and natural language processing (NLP) are rapidly evolving, applicability to harness large data to improve healthcare delivery is garnering great interest. Assessing antiepileptic drug (AED) efficacy remains a barrier to improving epilepsy care. In this review, we examined automatic electronic health record (EHR) extraction methodologies pertinent to epilepsy examining AED efficacy. We also reviewed more generalizable NLP pipelines to extract other critical patient variables.

Our review found varying reports of performance measures. Whereas automated data extraction pipelines are a crucial advancement, this review calls attention to standardizing NLP methodology and accuracy reporting for greater generalizability. Moreover, the use of crowdsourcing competitions to spur innovative NLP pipelines would further advance this field.

**HIGHLIGHTS:**

- Automated data extraction is rapidly evolving and can be harnessed to efficiently mine the electronic health record.
- Natural language processing (NLP) of unstructured text improves data extraction accuracy when added to ICD coding and structured fields.
- We review these techniques specific to epilepsy and highlight strengths as well as areas of further improvement.

## 1. BACKGROUND

### 1.1 Rationale

Epilepsy affects 60 million people worldwide.[1] Anti-epileptic drugs (AEDs) are first-line therapy for epilepsy and control seizures in two thirds of patients.[2] More than 25 AEDs are now available and rational “trial and error” often determines drug choice, as comparative data on efficacy between AEDs remains limited. Despite millions of people taking AEDs daily, retrospective and prospective chart review studies comparing AEDs head-to-head are only available for a limited number of medications. Moreover, small samples sizes limit interpretation of these studies. A wealth of information regarding AED efficacy lies within electronic health records (EHRs), yet efficient data extraction has been a critical barrier to closing this knowledge gap.

### 1.2 Overview

The goal of this report is to explore how pertinent data can be automatically extracted from the EHR for studies of AED efficacy, including comparative AED efficacy. The exposure variable is AED prescription. The outcome variable of seizure frequency can be assessed by comparing pre- and post-drug seizure frequencies. To compare two or more AEDs, cohorts of patients taking the AEDs of interest can be matched on relevant covariables to minimize confounding and relative changes in seizure frequency over time can be measured.

Patient characteristics that define the clinical context (Table 1) are important to include in the analysis as they have the potential to cause confounding effects in multiple ways. Demographic variables, such as socioeconomic status (SES), comorbidities (including psychiatric conditions such as substance abuse), and medication allergies may play a role in how providers choose which AED to trial, exacerbating nonrandom assignment to treatment groups. Underlying epilepsy etiology, baseline epilepsy severity, and age of onset may also correlate with refractoriness to AED treatment. Furthermore, underlying cause of epilepsy may have interaction effects with particular medications, such as the indication for broad-spectrum agents in the treatment of generalized epilepsy.

**TABLE 1:**
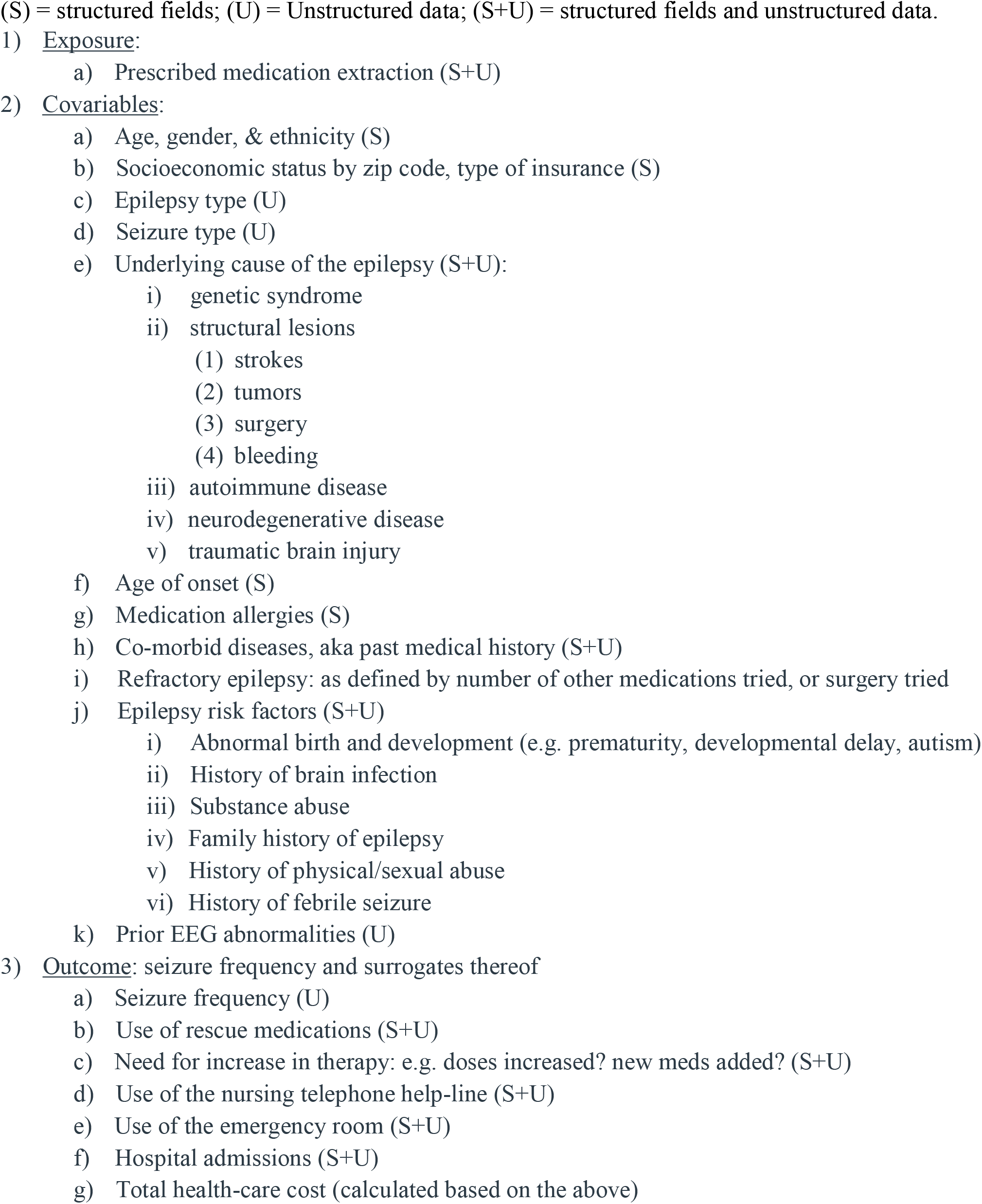
**KEY VARIABLES:** If these variables could be extracted from the electronic health record automatically, an antiepileptic drug efficacy study could be performed.

In this investigation, we performed a literature review of the currently available data extraction methods for the pertinent variables and examined techniques specific to epilepsy when appropriate to the variable (e.g., seizure frequency).[3] For more generalizable variables (e.g., medication), we also reviewed methods developed outside the epilepsy field.

### 1.3 Search strategy

We performed a PubMed search for the variables of interest listed in Table 1. The specific PubMed search phrases are described in detail in the supplementary text. Abstracts were screened for relevance, i.e. employment of a method of automated EHR extraction for one of the pertinent variables. Some manuscripts focused on the details of a method of extraction and its accuracy, while other manuscripts employed automated extraction methods as a means to describe a clinical outcome; both types of manuscripts were deemed relevant. When a manuscript was deemed relevant, we reviewed and catalogued elements of the manuscript including the summary of the extraction method, summary of accuracy, and estimate of generalizability to epilepsy as applicable (i.e. the method was not specifically created for epilepsy). Citations within relevant manuscripts were also reviewed in the same manner.

### 1.4 Terminology

Data in the EHR comprises both structured and unstructured fields. Structured data fields use controlled vocabulary and limit the variability, which allows more inter-user consistency and more accurate data aggregation. As examples of structured fields, blood pressure must be populated by exactly two numerical values (systolic and diastolic pressure), a medication field must be populated by a recognized medication name as selected from a standardized list, or a medical problem list must be populated by selecting from a list of Intelligent Medical Objects (IMOs) or International Statistical Classification of Disease codes (ICD codes).

Conversely, unstructured data components are composed of narrative text (prose) written by the provider, usually in the setting of a progress note in a clinic, emergency room, or inpatient encounter. Telephone calls are also often documented with free text. Natural Language Progressing (NLP) uses computer algorithms to extract information from unstructured free text language. Simple forms of NLP use dictionaries (lists of terms or synonyms) and rules (pre-set sentence structures) to extract information. More complex forms of NLP use machine learning to create a classifier which categorizes a note with the presence or absence of a particular variable. NLP algorithms and machine learning processes are often compiled into larger pipelines.

Measures of accuracy are critical to assess the performance of each method. In the field of data retrieval, the most commonly reported measures of accuracy are precision, recall, and the F1 statistic. Precision is the proportion of retrieved data that is true (positive predictive value). Recall is the proportion of true data that is correctly retrieved (sensitivity). The F1 statistic is the geometric mean between precision and recall; this statistic ranges from zero to one, where the value of one indicates perfect accuracy. Another summary measure that combines sensitivity and specificity is the area under the receiver operator curve (AUC); this statistic ranges from zero to one, where a value of one indicates perfect accuracy. For any extraction algorithm measuring a given variable, precision and recall can be reported on a training set (the same set of data that was used to create the extraction algorithm) or on a test set (an independent set of data that was not used to develop the extraction algorithm).

This report will reference specific EHR extraction algorithms and pipelines, including complex NLP machine learning methods (please see table 2), and report their measures of accuracy. In this review, all statistics were assumed to be reported on independent test sets unless specifically stated otherwise. Further elaboration of specific methodology of the pipelines is beyond the scope of this review.

**TABLE 2:**
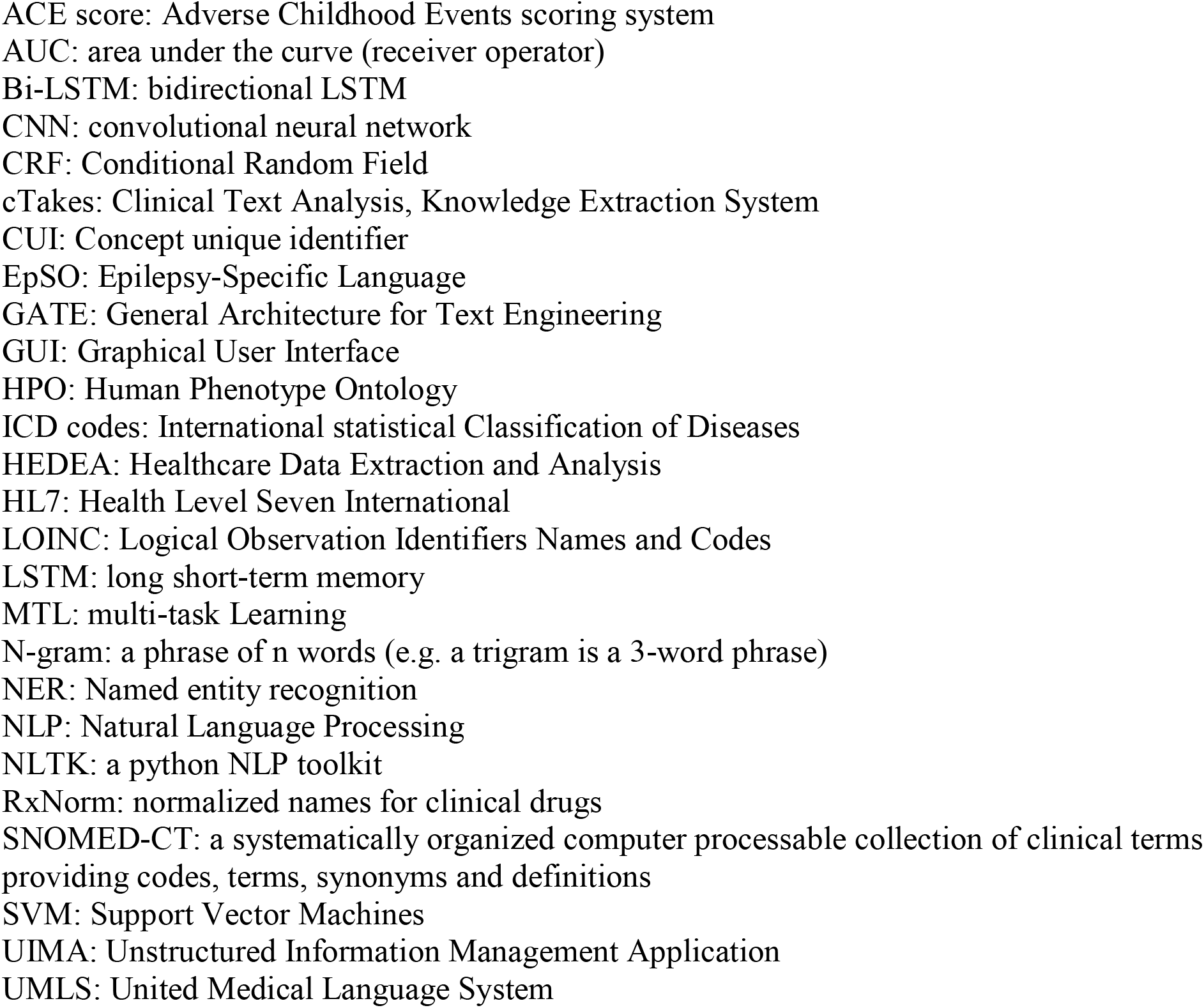
Acronyms and abbreviations for data standards and methods of machine learning

## 2. RESULTS

Over 2000 articles were returned by the PubMed search criteria and screened for relevance. A total of 128 articles were deemed sufficiently relevant for detailed review.

### 2.1 EXPOSURE VARIABLE

#### 2.1.1 Medication

There are a variety of emerging techniques to extract medications from the EHR, in part due to publicly issued crowdsource challenges which have generated a large number of highly accurate pipelines.[4] Extraction can be challenging as medications recorded within the EHR have variable interpretations depending on the context, as records may include current medications, past medications, recommended future medications, or medication allergies.[5] Many analysts suggest methods to account for this, such splitting a clinical note into various sections (i.e., the “current medications”, “allergies”, and “recommendations” sections).[6,7] The best approaches include analyzing a combination of structured fields and unstructured notes (e.g. as demonstrated by Cimino et al, though it must be noted that methods for unstructured notes have improved since this paper was published in 2007); this approach should account for potential differences in prescribing patterns among institutions and/or medication documentation in the EHR.[6,8]

The best pipeline for extraction of active medication ingredients and dosing achieves greater than 97% F1s.[9,10] However, extracting the indication for each medication is more difficult, with the best group achieving F1 of 66%.[11] Extracting the indication is particularly important in the field of epilepsy, since many of our medications may be prescribed for alternative indications (e.g. topiramate for migraine, oxcarbazepine for mood stabilization, etc.).

### 2.2 COVARIABLES

#### 2.2.1 Age and Sex

Age and binary sex are extracted reliably from structured fields.[12] Better methods are needed to account for nonbinary gender designations.

#### 2.2.2 Race and ethnicity

Race and ethnicity are often extracted from structured fields.[12–16] However, this method can be incompletely sensitive, and missing data tend to be biased, with underreporting of underrepresented socioeconomic demographics.[17–20] When ethnicity is present in structured fields, Denny et al report over 90% concordance with genetic ethnicity, which can be used as a gold standard for ethnicity extraction when available.[17,21] Sholle et al augmented structured fields with simple NLP to achieve an F1 of 91%.[19]

#### 2.2.3 Socioeconomic status (SES)

Some authors were able to extract values from structured fields while others used relatively simple forms of NLP to improve data or population statistics to extrapolate based on geography, such as the use of postal codes to impute SES.[22] Bejan et al captured “homelessness” with an AUC of 0.83.[23] Hatef et al showed that a simple NLP algorithm can supplement structured codes for “financial strain” and “housing issues” and increase recall by 10-15-fold over the use of ICD codes alone.[18] Biro et al leveraged Canadian census tract data by zip code using a combined deprivation index, which combines several ‘material’ and ‘social’ variables (such as income, education, living alone or with a spouse, etc.) to derive a single measure of SES.[24] Hollister et al used simple search terms to extract “low education” (PPV 80%), occupation (85%), unemployment (88%), retirement (64%), uninsured status (23%), Medicaid status (82%), and homelessness (33%).[25]

#### 2.2.4 Epilepsy type and seizure type

Focal epilepsy (FE), generalized epilepsy (GE), and unknown epilepsy (UE) can be discriminated by n-gram and an SVM classifier, as done by Connolly et al. These authors trained the model on data from one institution and tested on data from two others (F1 = 72%), which suggests reasonable generalizability; performance improved when trained on data from two institutions and tested on data from a third (F1 = 80%).[26] Cui et al defined a method PEEP (phenotype extraction in epilepsy) to extract epileptogenic zone, etiology, and EEG pattern from Epilepsy Monitoring Unit (EMU) discharge summaries. F1s ranged between 75% to 85% for exact matches for semiology, lateralizing signs, and EEG pattern, and up to 95% for epileptogenic zone. This method is the best method reviewed for specific semiology and etiologies but works on only EMU discharge summary note types.[27]

The most comprehensive pipeline specific to the field of epilepsy is ExECT (extraction of epilepsy clinical text).[28] This extracts a diagnosis of epilepsy as a binary field (88% precision, 89% recall), focal seizures as a binary field (96% precision, 70% recall), generalized seizures as a binary field (89% precision, 52% recall), and epilepsy type as a trinary field defined as either focal, generalized, or absence (90% precision, 80% recall).

#### 2.2.5 Underlying cause of epilepsy

A combination of approaches has been used which include extracting text from unstructured neuroradiology reports, structured comorbid disease fields, and the unstructured impression section of clinician notes. For certain specific etiologies for which no known automated EHR extraction exists (e.g. cortical dysplasia), raw image processing algorithms may be used. The ExECT pipeline identified abnormal or epileptogenic imaging findings on CT (56% precision, 59% recall) and MRI (82% precision, 69% recall) from clinical encounters, though these were limited to binary yes/no variables.[28] For brain tumors, Senders et al identified metastatic brain tumors from radiology notes, achieving AUC of 0.92 and best accuracy of 83%.[29]

For cortical dysplasia, Kassubek extracted information from raw imaging data, successfully identifying dysplasia locations in seven out of seven patients, using 30 controls as a reference.[30] For mesial temporal sclerosis, Chupin et al identified hippocampus and amygdala atrophy from raw imaging data to within approximately 10% of the “gold standard” based on structural volume.[31,32] For cerebral aneurysm, Castro et al improved on an initial screen of ICD codes with a simple dictionary and classifier NLP to achieve 86% PPV.[12] For venous thromboembolism, Heit et al combined ICD codes and NLP to achieve 100% PPV and 94% NPV.[21] Existing pipelines for flexible information extraction from free-text radiology reports have not been validated in epilepsy but may be repurposed for use in epilepsy.[33–35]

#### 2.2.6 Prior EEG abnormalities

In addition to direct analysis of the raw EEG signal, EEG abnormalities can be captured by means of structured fields and unstructured reports. Biswal et al achieved an AUC of 0.99 for detecting reports with seizures and AUC of 0.96 for epileptiform discharges, but a limitation of their study is that they did not differentiate focal from generalized findings.[36]

Bao reported 94% accuracy in interictal EEG diagnosis from raw EEG signal. However, this does not improve on the accuracy obtained from structured fields and unstructured reports, and increases the computational resources required for analysis as well as the administrative cost to obtain the raw EEG signal data.[37]

#### 2.2.7 Epilepsy severity

Epilepsy severity can be estimated by several characteristics which may be included in a model such as the number of current AEDs and historically prescribed AEDs (see the medications section above) or baseline seizure frequency (see seizure frequency section). Wissel et al used n-grams (up to n=3) and SVM to achieve sensitivity 80%, specificity 77%, PPV 25%, and NPV 98% for determining medically refractory epilepsy.[38]

#### 2.2.8 Age of onset

Extracting the age of onset in epilepsy remains an unmet challenge. Methods exist to extract the age of onset for other diseases in family history sections (i.e. age of cancer onset in family member).[39,40] However, these methods are not readily convertible to extract age of onset of a patient with epilepsy.

#### 2.2.9 Medication allergies and adverse drug effects

As noted in the medication section, prior studies reported that typical toolkits for NLP on clinical notes did not work well to abstract medications unless the note was split into sections.[41] The best available pipeline to identify medication allergies among those reviewed was that of Munkhdalai et who showed that an SVM model achieved the best average F1 of 89% on test data.[42]

#### 2.2.10 Comorbidities and past medical history

Comorbidities are often found in structured problem and diagnosis lists, but these are notoriously underpopulated by physicians, and thus need to be supplemented with free text extraction.[43,44]

Ning et al described SEDFE (SEmantics-Driven Feature Extraction) to collect medical concepts from online knowledge sources as candidate features and derived methods to achieve AUCs ranging from 0.90-0.95.[45] Other pipelines reviewed achieved slightly lower AUCs with widely differing methodologies,[46,47] including the repurposing of a crowdsourcing marketplace.[48]

Capturing common psychiatric comorbidities associated epilepsy remain challenging. Commonly, comorbidities such as anxiety and depression can be captured as a diagnosis or solely as patient-reported symptoms. Furthermore, these comorbidities are likely underdiagnosed and undertreated within the epilepsy population. Validated depression instruments, such as the Patient Health Questionnarie-9 (PHQ-9) to assess depressive symptoms, can be easily extracted from structured fields. However, Adekkanattu et al implemented an NLP platform to extract PHQ-9 scoring from unstructured clinical text for patients prescribed an antidepressant with high accuracy (F-score 97%) and found that nearly one-third of patients’ charts had a score that clinically indicated major depressive disorder without a structured ICD diagnosis code associated.[49] In a study to predict advanced care of depression via statewide EHR data, free text extraction from decision tree models yielded AUC scores of approximately 90% for patients deemed high-risk patients versus approximately 80% for the overall patient population, respectively.[50]

#### 2.2.11 Epilepsy risk factors – history of drug abuse

The main work in this area has been developed for the detection of opioid dependence and smoking.[51] Notably, many authors have reported that ICD codes are insufficient for the accurate diagnosis of opioid dependence or overdose.[52–54] NLP can help improve recall of drug abuse from unstructured fields in clinician’s notes.[55–57] Including nursing notes further improves performance.[58] Hazlehurst et al demonstrated that NLP can be generalizable by publishing the difference in accuracy between their training and test sets.[54]

Variables more applicable to seizure risk include alcohol abuse and stimulant abuse. [58–61] These variables have not been studied in as much detail as opiates and nicotine. The most accurate study we reviewed is that of Wang et al which attained F1s of 90% for alcohol abuse and 85% for drug abuse detection.[60]

#### 2.2.12 Epilepsy risk factors – family history of epilepsy

Family history statements have been extracted from a variety of note types for a variety of diseases, [62–66] and it is likely that these can be repurposed to epilepsy. Zhou et al were able to achieve precision of 100% and recall of 97% using NLP, but their method is limited to looking at discharge summaries and admission notes.[65,66] Mowery et al achieved precision 96% and recall 94% using NLP on clinicians’ notes.[40]

### 2.3 OUTCOME VARIABLES

#### 2.3.1 Primary Outcome - Seizure frequency

The ExECT pipeline (methods discussed in Epilepsy Type section above) identified the phrase or sentence within a clinical document that contained the seizure frequency but does not return a numeric value, with precision 86% and recall 54%.[28] To our knowledge, this is the only study of seizure frequency in the literature.

#### 2.3.2 Secondary Outcomes- Use of rescue medications and the total number of AEDs required

Recurrent seizures may be identified by the use of rescue medications or the need to add additional AEDs. See the medication section above for details on how medications can be extracted.

### 2.4 OTHER GENERAL USE PIPELINES

Our PubMed search results also found a series of general use pipelines, documented in detail in the supplementary text. One effort worth noting is that Kannan et al propose a strategy for incorporating prospective research-quality data collection into the practitioners’ workflow without burdening practitioners with excessive documentation, which is the primary barrier to this type of collection.[67,68] They were able to implement prospective cohort building pipelines in 43 chronic diseases.[67] The implementation of such a framework within the field of epilepsy would allow for prospective data collection which is known to be superior to retrospective studies.

### 2.5 PIPELINES IN OTHER DIALECTS AND LANGUAGES

For greater applicability and sample size, a multi-center international study would require the use of pipelines in many languages. As of now, there appears to be minimal ability to convert pipelines from one language to another. There is one notable study where NLP developed in Europe and the United States was applied to medical notes written in Indian-English to extrac medical diagnoses, labs, procedures, demographic information, and outcomes.[69] Details about pipelines in other languages can be found in the supplementary material.

## 3. SUMMARY AND CONCLUSION

Automated extraction of EHR data has advanced impressively in the last several years, with a plethora of methods published to date. However, there is great variability in algorithm performance across variables. We have seen that some variables can be easily and accurately extracted from the EHR by way of structured and unstructured fields, such as age, sex, and family history. Other variables can be extracted with reasonably high sensitivity but with lower specificity, for example, SES, ethnicity, epilepsy risk factors, and EEG and MRI results.

Medications can be extracted with high accuracy (precision and recall > 95%) using a number of approaches, thanks in part due to public challenges that awarded prize money to competing teams to crowdsource the best method. Furthermore, techniques that are developed in fields outside of epilepsy can easily be applied to epilepsy.

Several variables cannot be reliably extracted with current published methods, including seizure etiology and epilepsy severity. The problem of seizure etiology may potentially be solved by a multi-modal approach incorporating EEG findings, MRI findings, structured and unstructured fields, and setting up decision trees based on ILAE diagnostic criteria. This approach would be relatively easy to implement if the underlying variables could be extracted reliably (EEG findings, MRI findings, and comorbidities). Epilepsy severity is not a single variable, but rather a concept which requires the incorporation of several variables including the number of medications used, the seizure frequency, presence of convulsive seizures, and potentially electroencephalographic markers.

The greatest limitation to assessing AED efficacy using EHR data is that the outcome of primary interest, seizure frequency, cannot be extracted reliably from the record using currently available techniques. The best techniques can identify only the seizure frequency text with poor sensitivity (∼50%) and only marginal specificity (∼80%). Future approaches to increase sensitivity will undoubtedly come at a cost of specificity. However, one could envision an approach to create semi-automated algorithms, wherein an automated method screens notes comprehensively for key sentences with subsequent manual review to extract seizure frequency quantitatively and with greater accuracy. The limitation of a semi-automated method would be the time required for human review of each data-reduced chart. Within the NLP field, the continued use of crowdsourcing will be vital to creating new pipelines and increasing accuracy to optimize data extraction from EHRs. Notably, medication extraction, medication allergy extraction, and opioid use and dependence can all be readily and accurately extracted with one of several available pipelines produced through crowdsourcing competitions.

As automated extraction methods continue to evolve, standards on reporting the accuracy of these pipelines should be followed. This will allow for comparisons to be drawn between methods. We call for, at minimum, the reporting of precision, recall, and F1 statistics for training and test sets (when a test set is available). We also recommend for all studies to use an independent test set when possible, and ideally another independent validation dataset from a different institution. This reporting is crucial because the decrease in accuracy between the training set and the test set is informative of the algorithm’s generalizability. We also appreciate that most of the studies that we reviewed have provided public access to the extraction algorithms, and we encourage all authors to do the same.

In summary, we evaluated the feasibility, availability, and performance of automated data extraction methods to facilitate prospective and retrospective investigation of AED efficacy. The most significant roadblock is the derth of algorithms to extract seizure frequency. Other smaller but important roadblocks are the extraction of seizure etiology and epilepsy severity.

## Data Availability

The authors confirm that the data supporting the findings of this study are available within the article and its supplementary materials.

## ACKNOWLEDGEMENTS

The authors would like to acknowledge Brian Litt, MD for his expertise and support.

## REFERENCES

[1] Allers K, Essue BM, Hackett ML, Muhunthan J, Anderson CS, Pickles K, et al. The economic impact of epilepsy: a systematic review. BMC Neurol 2015;15:245. https://doi.org/10.1186/s12883-015-0494-y.

[2] Eatock J, Baker GA. Managing patient adherence and quality of life in epilepsy. Neuropsychiatr Dis Treat 2007;3:117–31. https://doi.org/10.2147/nedt.2007.3.1.117.

[3] Velupillai S, Suominen H, Liakata M, Roberts A, Shah AD, Morley K, et al. Using clinical Natural Language Processing for health outcomes research: Overview and actionable suggestions for future advances. J Biomed Inform 2018;88:11–9. https://doi.org/10.1016/j.jbi.2018.10.005.

[4] Jagannatha A, Liu F, Liu W, Yu H. Overview of the First Natural Language Processing Challenge for Extracting Medication, Indication, and Adverse Drug Events from Electronic Health Record Notes (MADE 1.0). Drug Saf 2019;42:99–111. https://doi.org/10.1007/s40264-018-0762-z.

[5] Chhieng D, Day T, Gordon G, Hicks J. Use of natural language programming to extract medication from unstructured electronic medical records. AMIA. Annu Symp Proceedings AMIA Symp 2007:908.

[6] Sohn S, Clark C, Halgrim SR, Murphy SP, Jonnalagadda SR, Wagholikar KB, et al. Analysis of Cross-Institutional Medication Description Patterns in Clinical Narratives. Biomed Inform Insights 2013;6s1:BII.S11634. https://doi.org/10.4137/BII.S11634.

[7] Farooq F, Yu S, Anand V, Krishnapuram B. Categorizing medications from unstructured clinical notes. AMIA Jt Summits Transl Sci Proceedings AMIA Jt Summits Transl Sci 2013;2013:48–52.

[8] Cimino JJ, Bright TJ, Li J. Medication reconciliation using natural language processing and controlled terminologies. Stud Health Technol Inform 2007;129:679–83.

[9] Dietrich G, Krebs J, Liman L, Fette G, Ertl M, Kaspar M, et al. Replicating medication trend studies using ad hoc information extraction in a clinical data warehouse. BMC Med Inform Decis Mak 2019;19:15. https://doi.org/10.1186/s12911-018-0729-0.

[10] Jiang M, Wu Y, Shah A, Priyanka P, Denny JC, Xu H. Extracting and standardizing medication information in clinical text - the MedEx-UIMA system. AMIA Jt Summits Transl Sci Proceedings AMIA Jt Summits Transl Sci 2014;2014:37–42.

[11] Li F, Liu W, Yu H. Extraction of Information Related to Adverse Drug Events from Electronic Health Record Notes: Design of an End-to-End Model Based on Deep Learning. JMIR Med Informatics 2018;6:e12159. https://doi.org/10.2196/12159.

[12] Castro VM, Dligach D, Finan S, Yu S, Can A, Abd-El-Barr M, et al. Large-scale identification of patients with cerebral aneurysms using natural language processing. Neurology 2017;88:164–8. https://doi.org/10.1212/WNL.0000000000003490.

[13] Gundlapalli A V., Jones AL, Redd A, Divita G, Brignone E, Pettey WBP, et al. Combining Natural Language Processing of Electronic Medical Notes With Administrative Data to Determine Racial/Ethnic Differences in the Disclosure and Documentation of Military Sexual Trauma in Veterans. Med Care 2019;57:S149–56. https://doi.org/10.1097/MLR.0000000000001031.

[14] Kaur H, Sohn S, Wi C-I, Ryu E, Park MA, Bachman K, et al. Automated chart review utilizing natural language processing algorithm for asthma predictive index. BMC Pulm Med 2018;18:34. https://doi.org/10.1186/s12890-018-0593-9.

[15] Meystre SM, Thibault J, Shen S, Hurdle JF, South BR. Textractor: a hybrid system for medications and reason for their prescription extraction from clinical text documents. J Am Med Informatics Assoc 2010;17:559–62. https://doi.org/10.1136/jamia.2010.004028.

[16] Zheng C, Luo Y, Mercado C, Sy L, Jacobsen SJ, Ackerson B, et al. Using natural language processing for identification of herpes zoster ophthalmicus cases to support population-based study. Clin Experiment Ophthalmol 2019;47:7–14. https://doi.org/10.1111/ceo.13340.

[17] Denny JC. Chapter 13: Mining Electronic Health Records in the Genomics Era. PLoS Comput Biol 2012;8:e1002823. https://doi.org/10.1371/journal.pcbi.1002823.

[18] Hatef E, Rouhizadeh M, Tia I, Lasser E, Hill-Briggs F, Marsteller J, et al. Assessing the Availability of Data on Social and Behavioral Determinants in Structured and Unstructured Electronic Health Records: A Retrospective Analysis of a Multilevel Health Care System. JMIR Med Informatics 2019;7:e13802. https://doi.org/10.2196/13802.

[19] Sholle ET, Pinheiro LC, Adekkanattu P, Davila MA, Johnson SB, Pathak J, et al. Underserved populations with missing race ethnicity data differ significantly from those with structured race/ethnicity documentation. J Am Med Informatics Assoc 2019;26:722–9. https://doi.org/10.1093/jamia/ocz040.

[20] Wissel BD, Greiner HM, Glauser TA, Mangano FT, Santel D, Pestian JP, et al. Investigation of bias in an epilepsy machine learning algorithm trained on physician notes. Epilepsia 2019;60:e93–8. https://doi.org/10.1111/epi.16320.

[21] Heit JA, Armasu S, McCauley B, Kullo I, Sicotte H, Pathak J, et al. Identification of unique venous thromboembolism-susceptibility variants in African-Americans. Thromb Haemost 2017;117:758–68. https://doi.org/10.1160/TH16-08-0652.

[22] Dandona L, Dandona R, Naduvilath TJ, McCarty CA, Mandal P, Srinivas M, et al. Population-based assessment of the outcome of cataract surgery in an urban population in southern India. Am J Ophthalmol 1999;127:650–8. https://doi.org/10.1016/s0002-9394(99)00044-6.

[23] Bejan CA, Angiolillo J, Conway D, Nash R, Shirey-Rice JK, Lipworth L, et al. Mining 100 million notes to find homelessness and adverse childhood experiences: 2 case studies of rare and severe social determinants of health in electronic health records. J Am Med Informatics Assoc 2018;25:61–71. https://doi.org/10.1093/jamia/ocx059.

[24] Biro S, Williamson T, Leggett JA, Barber D, Morkem R, Moore K, et al. Utility of linking primary care electronic medical records with Canadian census data to study the determinants of chronic disease: an example based on socioeconomic status and obesity. BMC Med Inform Decis Mak 2016;16:32. https://doi.org/10.1186/s12911-016-0272-9.

[25] Hollister BM, Restrepo NA, Farber-Eger E, Crawford DC, Aldrich MC, Non A. DEVELOPMENT AND PERFORMANCE OF TEXT-MINING ALGORITHMS TO EXTRACT SOCIOECONOMIC STATUS FROM DE-IDENTIFIED ELECTRONIC HEALTH RECORDS. Biocomput. 2017, WORLD SCIENTIFIC; 2017, p. 230–41. https://doi.org/10.1142/9789813207813_0023.

[26] Connolly B, Matykiewicz P, Bretonnel Cohen K, Standridge SM, Glauser TA, Dlugos DJ, et al. Assessing the similarity of surface linguistic features related to epilepsy across pediatric hospitals. J Am Med Informatics Assoc 2014;21:866–70. https://doi.org/10.1136/amiajnl-2013-002601.

[27] Cui L, Sahoo SS, Lhatoo SD, Garg G, Rai P, Bozorgi A, et al. Complex epilepsy phenotype extraction from narrative clinical discharge summaries. J Biomed Inform 2014;51:272–9. https://doi.org/10.1016/j.jbi.2014.06.006.

[28] Fonferko-Shadrach B, Lacey AS, Roberts A, Akbari A, Thompson S, Ford D V, et al. Using natural language processing to extract structured epilepsy data from unstructured clinic letters: development and validation of the ExECT (extraction of epilepsy clinical text) system. BMJ Open 2019;9:e023232. https://doi.org/10.1136/bmjopen-2018-023232.

[29] Senders JT, Karhade A V., Cote DJ, Mehrtash A, Lamba N, DiRisio A, et al. Natural Language Processing for Automated Quantification of Brain Metastases Reported in Free-Text Radiology Reports. JCO Clin Cancer Informatics 2019:1–9. https://doi.org/10.1200/CCI.18.00138.

[30] Kassubek J, Huppertz H-J, Spreer J, Schulze-Bonhage A. Detection and localization of focal cortical dysplasia by voxel-based 3-D MRI analysis. Epilepsia 2002;43:596–602. https://doi.org/10.1046/j.1528-1157.2002.41401.x.

[31] Chupin M, Mukuna-Bantumbakulu AR, Hasboun D, Bardinet E, Baillet S, Kinkingnéhun S, et al. Anatomically constrained region deformation for the automated segmentation of the hippocampus and the amygdala: Method and validation on controls and patients with Alzheimer’s disease. Neuroimage 2007;34:996–1019. https://doi.org/10.1016/j.neuroimage.2006.10.035.

[32] Istephan S, Siadat M-R. Unstructured medical image query using big data – An epilepsy case study. J Biomed Inform 2016;59:218–26. https://doi.org/10.1016/j.jbi.2015.12.005.

[33] Pons E, Braun LMM, Hunink MGM, Kors JA. Natural language processing in radiology: A systematic review. Radiology 2016;279:329–43. https://doi.org/10.1148/radiol.16142770.

[34] Hassanpour S, Langlotz CP. Information extraction from multi-institutional radiology reports. Artif Intell Med 2016;66:29–39. https://doi.org/10.1016/j.artmed.2015.09.007.

[35] Steinkamp JM, Chambers C, Lalevic D, Zafar HM, Cook TS. Toward Complete Structured Information Extraction from Radiology Reports Using Machine Learning. J Digit Imaging 2019. https://doi.org/10.1007/s10278-019-00234-y.

[36] Biswal S, Nip Z, Moura Junior V, Bianchi MT, Rosenthal ES, Westover MB. Automated information extraction from free-text EEG reports. 2015 37th Annu. Int. Conf. IEEE Eng. Med. Biol. Soc., IEEE; 2015, p. 6804–7. https://doi.org/10.1109/EMBC.2015.7319956.

[37] Forrest Sheng Bao, Jue-Ming Gao, Jing Hu, Donald Lie, Yuanlin Zhang, Oommen KJ. Automated epilepsy diagnosis using interictal scalp EEG. 2009 Annu. Int. Conf. IEEE Eng. Med. Biol. Soc., IEEE; 2009, p. 6603–7. https://doi.org/10.1109/IEMBS.2009.5332550.

[38] Wissel BD, Greiner HM, Glauser TA, Holland-Bouley KD, Mangano FT, Santel D, et al. Prospective validation of a machine learning model that uses provider notes to identify candidates for resective epilepsy surgery. Epilepsia 2020;61:39–48. https://doi.org/10.1111/epi.16398.

[39] Sin M, McGuinness JE, Trivedi MS, Vanegas A, Silverman TB, Crew KD, et al. Automatic Genetic Risk Assessment Calculation Using Breast Cancer Family History Data from the EHR compared to Self-Report. AMIA. Annu Symp Proceedings AMIA Symp 2018;2018:970–8.

[40] Mowery DL, Kawamoto K, Bradshaw R, Kohlmann W, Schiffman JD, Weir C, et al. Determining Onset for Familial Breast and Colorectal Cancer from Family History Comments in the Electronic Health Record. AMIA Jt Summits Transl Sci Proceedings AMIA Jt Summits Transl Sci 2019;2019:173–81.

[41] Chen L, Gu Y, Ji X, Sun Z, Li H, Gao Y, et al. Extracting medications and associated adverse drug events using a natural language processing system combining knowledge base and deep learning. J Am Med Informatics Assoc 2020;27:56–64. https://doi.org/10.1093/jamia/ocz141.

[42] Munkhdalai T, Liu F, Yu H. Clinical Relation Extraction Toward Drug Safety Surveillance Using Electronic Health Record Narratives: Classical Learning Versus Deep Learning. JMIR Public Heal Surveill 2018;4:e29. https://doi.org/10.2196/publichealth.9361.

[43] Karmakar C, Luo W, Tran T, Berk M, Venkatesh S. Predicting Risk of Suicide Attempt Using History of Physical Illnesses From Electronic Medical Records. JMIR Ment Heal 2016;3:e19. https://doi.org/10.2196/mental.5475.

[44] Sheehan OC, Kharrazi H, Carl KJ, Leff B, Wolff JL, Roth DL, et al. Helping Older Adults Improve Their Medication Experience (HOME) by Addressing Medication Regimen Complexity in Home Healthcare. Home Healthc Now 2018;36:10–9. https://doi.org/10.1097/NHH.0000000000000632.

[45] Ning W, Chan S, Beam A, Yu M, Geva A, Liao K, et al. Feature extraction for phenotyping from semantic and knowledge resources. J Biomed Inform 2019;91:103122. https://doi.org/10.1016/j.jbi.2019.103122.

[46] Chen Q, Li H, Tang B, Wang X, Liu X, Liu Z, et al. An automatic system to identify heart disease risk factors in clinical texts over time. J Biomed Inform 2015;58:S158–63. https://doi.org/10.1016/j.jbi.2015.09.002.

[47] Gronsbell J, Minnier J, Yu S, Liao K, Cai T. Automated feature selection of predictors in electronic medical records data. Biometrics 2019;75:268–77. https://doi.org/10.1111/biom.12987.

[48] Yetisgen-Yildiz M, Solti I, Xia F. Using Amazon’s Mechanical Turk for Annotating Medical Named Entities. AMIA. Annu Symp Proceedings AMIA Symp 2010;2010:1316.

[49] P A, ET S, J D, J P, SB J, TR C. Ascertaining Depression Severity by Extracting Patient Health Questionnaire-9 (PHQ-9) Scores From Clinical Notes. AMIA. Annu Symp Proceedings AMIA Symp 2018;2018.

[50] Kasthurirathne SN, Biondich PG, Grannis SJ, Purkayastha S, Vest JR, Jones JF. Identification of Patients in Need of Advanced Care for Depression Using Data Extracted From a Statewide Health Information Exchange: A Machine Learning Approach. J Med Internet Res 2019;21:e13809. https://doi.org/10.2196/13809.

[51] Uzuner O, Goldstein I, Luo Y, Kohane I. Identifying patient smoking status from medical discharge records. J Am Med Inform Assoc 2008;15:14–24. https://doi.org/10.1197/jamia.M2408.

[52] Afshar M, Joyce C, Dligach D, Sharma B, Kania R, Xie M, et al. Subtypes in patients with opioid misuse: A prognostic enrichment strategy using electronic health record data in hospitalized patients. PLoS One 2019;14:e0219717. https://doi.org/10.1371/journal.pone.0219717.

[53] Haller I V., Renier CM, Juusola M, Hitz P, Steffen W, Asmus MJ, et al. Enhancing Risk Assessment in Patients Receiving Chronic Opioid Analgesic Therapy Using Natural Language Processing. Pain Med 2016:pnw283. https://doi.org/10.1093/pm/pnw283.

[54] Hazlehurst B, Green CA, Perrin NA, Brandes J, Carrell DS, Baer A, et al. Using natural language processing of clinical text to enhance identification of opioid-related overdoses in electronic health records data. Pharmacoepidemiol Drug Saf 2019;28:1143–51. https://doi.org/10.1002/pds.4810.

[55] Carrell DS, Cronkite D, Palmer RE, Saunders K, Gross DE, Masters ET, et al. Using natural language processing to identify problem usage of prescription opioids. Int J Med Inform 2015;84:1057–64. https://doi.org/10.1016/j.ijmedinf.2015.09.002.

[56] Palmer RE, Carrell DS, Cronkite D, Saunders K, Gross DE, Masters E, et al. The prevalence of problem opioid use in patients receiving chronic opioid therapy. Pain 2015;156:1208–14. https://doi.org/10.1097/j.pain.0000000000000145.

[57] Green CA, Perrin NA, Hazlehurst B, Janoff SL, DeVeaugh-Geiss A, Carrell DS, et al. Identifying and classifying opioid-related overdoses: A validation study. Pharmacoepidemiol Drug Saf 2019;28:1127–37. https://doi.org/10.1002/pds.4772.

[58] Topaz M, Murga L, Bar-Bachar O, Cato K, Collins S. Extracting Alcohol and Substance Abuse Status from Clinical Notes: The Added Value of Nursing Data. Stud Health Technol Inform 2019;264:1056–60. https://doi.org/10.3233/SHTI190386.

[59] Lingeman JM, Wang P, Becker W, Yu H. Detecting Opioid-Related Aberrant Behavior using Natural Language Processing. AMIA. Annu Symp Proceedings AMIA Symp 2017;2017:1179–85.

[60] Wang Y, Chen ES, Pakhomov S, Arsoniadis E, Carter EW, Lindemann E, et al. Automated Extraction of Substance Use Information from Clinical Texts. AMIA. Annu Symp Proceedings AMIA Symp 2015;2015:2121–30.

[61] Afshar M, Phillips A, Karnik N, Mueller J, To D, Gonzalez R, et al. Natural language processing and machine learning to identify alcohol misuse from the electronic health record in trauma patients: development and internal validation n.d. https://doi.org/10.1093/jamia/ocy166.

[62] Bill R, Pakhomov S, Chen ES, Winden TJ, Carter EW, Melton GB. Automated extraction of family history information from clinical notes. AMIA. Annu Symp Proceedings AMIA Symp 2014;2014:1709–17.

[63] Mehrabi S, Krishnan A, Roch AM, Schmidt H, Li D, Kesterson J, et al. Identification of Patients with Family History of Pancreatic Cancer--Investigation of an NLP System Portability. Stud Health Technol Inform 2015;216:604–8.

[64] Friedlin J, McDonald CJ. Using a natural language processing system to extract and code family history data from admission reports. AMIA. Annu Symp Proceedings AMIA Symp 2006;2006:925.

[65] Goss FR, Plasek JM, Lau JJ, Seger DL, Chang FY, Zhou L. An evaluation of a natural language processing tool for identifying and encoding allergy information in emergency department clinical notes. AMIA. Annu Symp Proceedings AMIA Symp 2014;2014:580–8.

[66] Zhou L, Plasek JM, Mahoney LM, Chang FY, DiMaggio D, Rocha RA. Mapping Partners Master Drug Dictionary to RxNorm using an NLP-based approach. J Biomed Inform 2012;45:626–33. https://doi.org/10.1016/j.jbi.2011.11.006.

[67] Kannan V, Fish J, Mutz J, Carrington A, Lai K, Davis L, et al. Rapid Development of Specialty Population Registries and Quality Measures from Electronic Health Record Data. Methods Inf Med 2017;56:e74–83. https://doi.org/10.3414/ME16-02-0031.

[68] Warner JL, Anick P, Hong P, Xue N. Natural Language Processing and the Oncologic History: Is There a Match? J Oncol Pract 2011;7:e15–9. https://doi.org/10.1200/JOP.2011.000240.

[69] Ramanan S V., Radhakrishna K, Waghmare A, Raj T, Nathan SP, Sreerama SM, et al. Dense Annotation of Free-Text Critical Care Discharge Summaries from an Indian Hospital and Associated Performance of a Clinical NLP Annotator. J Med Syst 2016;40:187. https://doi.org/10.1007/s10916-016-0541-2.

